# The Impact Mechanism of Screen Time on Depression Among Chinese College Students: A Chain Mediation Model of Sleep Quality and Emotion Regulation

**DOI:** 10.64898/2026.07.20.26358281

**Authors:** Liang Chen, Ding-you Zhang, Kai-xin Li, Bo Li, Hu Lou, Sai Zhu, Shi-hai Yu, Shan-shan Han

## Abstract

**Purpose:** This study aimed to examine the association between screen time and depressive symptoms among Chinese college students, and to investigate the mediating roles of sleep quality and emotion regulation in this relationship. Furthermore, a serial mediation model was constructed to elucidate the underlying psychological mechanisms linking screen exposure to depression.

**Methods:** A stratified cluster sampling method was employed to recruit 10,999 college students for a cross-sectional questionnaire survey. Data were collected on screen time, sleep quality, emotion regulation ability, and depressive symptoms. Descriptive statistics, correlation analyses, and regression analyses were conducted using SPSS 26.0 A serial mediation model was tested using the PROCESS macro (Model 6), and bootstrapping procedures were applied to estimate the significance of indirect effects.

**Results:** Correlation analyses indicated that screen time was significantly positively associated with depressive symptoms (r = 0.16, p < 0.01) and sleep quality (r = 0.15, p < 0.01), and significantly negatively associated with emotion regulation (r = −0.13, p < 0.01). Sleep quality was positively correlated with depressive symptoms (r = 0.31, p < 0.01), whereas emotion regulation was negatively correlated with depressive symptoms (r = −0.42, p < 0.01).

Regression analyses further showed that screen time significantly positively predicted depressive symptoms (β = 0.712, p < 0.001), positively predicted sleep quality (β = 0.217, p < 0.001), and negatively predicted emotion regulation (β = −0.085, p < 0.001). In addition, both sleep quality (β = 1.318, p < 0.001) and emotion regulation (β = −0.424, p < 0.001) were significant predictors of depressive symptoms.

Mediation analyses demonstrated that sleep quality significantly mediated the association between screen time and depressive symptoms (95% CI [0.239, 0.332]), as did emotion regulation (95% CI [0.269, 0.416]). Moreover, a significant serial mediation effect of sleep quality and emotion regulation was observed in the relationship between screen time and depressive symptoms (95% CI [0.082, 0.117]).

**Conclusion:** Screen time is significantly associated with depressive symptoms among college students, with sleep quality and emotion regulation serving as important mediating mechanisms. Extended screen exposure may be linked to higher levels of depressive symptoms by impairing sleep quality and weakening emotion regulation capacity.

## 0 Introduction

With the rapid proliferation of mobile internet, online learning platforms, and social media, digital screens have become deeply embedded in university students’ academic, social, and recreational lives. Screen time generally refers to the amount of time individuals spend using electronic screen-based devices such as smartphones, computers, tablets, and televisions, encompassing both academic and non-academic (e.g., entertainment and social) use. For university students, moderate screen use facilitates information acquisition, academic task completion, and social connectedness. However, excessive screen exposure may displace time allocated to sleep, physical activity, and face-to-face social interaction, thereby exerting adverse effects on physical and mental health [1]. In recent years, Chinese college students have demonstrated increasingly high levels of daily screen time, with smartphone use accounting for the largest proportion [2]. Given that academic learning, social communication, and entertainment activities are all highly dependent on digital devices, screen use has evolved from a simple leisure behavior into an integrated daily behavioral pattern [3]. Prolonged screen exposure has been associated with a range of physical and psychological problems. Among these, depression has attracted particular attention due to its profound impact on academic performance, interpersonal functioning, and suicide risk [4], making it one of the most critical issues in university students’ mental health research [5]. Depression is characterized by persistent low mood, diminished interest or pleasure, negative cognitive bias, and impaired social functioning [6]. For university students, depressive symptoms not only undermine academic engagement and performance [7], but also impair interpersonal relationships [8], life adaptation, and confidence in future development [9], As a pivotal group transitioning from adolescence to adulthood, the mental health status of college students is closely linked not only to individual academic and occupational outcomes, but also to the broader quality of future societal human capital and social stability. Therefore, identifying the determinants of depression and elucidating its underlying mechanisms among college students holds substantial theoretical and practical significance.

To systematically explain the underlying mechanisms linking screen time and depression among college students, this study integrates Media Displacement Theory (MDT), the Sleep–Emotion Regulation model (SQ-ER), and the Process Model of Emotion Regulation (PMER), and proposes a serial mediation framework characterized by “behavioral displacement → impaired physiological recovery → weakened psychological regulation.”

MDT posits that increased time spent on screen-based media inevitably displaces time allocated to traditional activities such as sleep, physical activity, and face-to-face social interaction[10]. Sleep serves as the foundation of physiological recovery, physical activity functions as a channel for stress relief [11], and real-world social interaction represents a core source of social support [12]. Accordingly, MDT provides the initial theoretical premise of this study: increased screen time may first undermine sleep resources through a time-displacement mechanism. However, MDT primarily addresses behavioral time allocation and does not explain how reduced sleep subsequently translates into mental health outcomes. To address this gap, the SQ-ER model is introduced as a second-layer explanatory mechanism. This model emphasizes that sleep is not only essential for physiological restoration but also critical for maintaining emotional stability and cognitive control [13]. Chronic sleep deprivation or poor sleep quality significantly impairs prefrontal cortex–mediated executive functioning, emotional recovery, and stress regulation, thereby increasing susceptibility to persistent negative affect [14]. Building on MDT, SQ-ER further clarifies how behavioral displacement gives rise to psychological consequences. Once sleep quality is compromised, a key question becomes how individuals regulate the resulting negative emotions—this process represents a critical juncture determining whether such emotional disturbances develop into clinically relevant depressive symptoms. The Process Model of Emotion Regulation (PMER) provides the third-layer explanatory mechanism. This model emphasizes that individuals regulate emotional states through strategies such as cognitive reappraisal and expressive suppression, which differentially influence mental health outcomes [15]. Prolonged sleep disturbance weakens the neurocognitive capacity required for adaptive strategies such as cognitive reappraisal [16]. Reduced emotion regulation capacity, in turn, increases reliance on maladaptive strategies, including avoidance, suppression, and rumination, thereby elevating the risk of depression.

Screen time refers to the amount of time individuals spend using electronic devices such as smartphones, computers, tablets, and televisions [17], typically encompassing both academic and entertainment-related use[18]. Moderate screen use can meet needs related to learning, information acquisition, and social communication [19], whereas excessive use may displace physical activity, face-to-face interaction, and sleep [20]. Empirical evidence consistently shows that Chinese college students spend more than six hours per day on screens, with smartphones accounting for approximately 50%–70% of total screen exposure [21, 22]. Theoretically, excessive screen time may increase depression risk through multiple pathways. First, greater time allocated to screen-based activities reduces time available for sleep, physical activity, and real-world social interaction, all of which serve protective psychological functions [23]. Second, prolonged exposure to social comparison, negative content, and information overload on digital platforms may foster maladaptive cognitive biases, thereby increasing depressive vulnerability [24]. Accordingly, this study proposes Hypothesis 1: screen time positively predicts depression among college students.

Sleep quality is a key indicator of sleep health, typically encompassing sleep latency, duration, efficiency, disturbances, and daytime functioning [25]. Due to academic demands, social engagement, employment pressure, and online entertainment, college students are particularly vulnerable to late bedtime, insufficient sleep, and poor sleep quality [26]. Excessive screen use—especially before bedtime—may delay sleep onset and disrupt circadian rhythms through blue light exposure, cognitive and emotional arousal, information overload, and immersive engagement [27]. Previous studies have demonstrated that screen exposure duration is associated with depressive symptoms among Chinese college students [28], and that sleep quality mediates the relationship between screen time and negative affect, suggesting that sleep is a key pathway linking screen exposure to depression, anxiety, and stress-related outcomes [29]. The World Health Organization has also noted that problematic social media use is associated with poorer mental and social well-being, later sleep onset, and sleep deprivation, which in turn constitute significant risk factors for depressive symptoms[30], According to the SQ-ER model, sleep is not only a physiological recovery process but also a foundational mechanism for emotional stability and cognitive control. When individuals experience chronic sleep deprivation or poor sleep quality, their emotional recovery capacity, stress coping ability, and cognitive regulation processes are significantly compromised, thereby increasing persistent negative affect[31], Accordingly, sleep quality may serve as a key mediating mechanism linking screen time and depression. Thus, Hypothesis 2 proposes that sleep quality mediates the relationship between screen time and depression.

Emotion regulation refers to the ability to identify, understand, express, and modulate one’s emotional experiences [32]. For college students, effective emotion regulation enables adaptive coping with academic stress, interpersonal conflict, employment anxiety, and life transitions through strategies such as cognitive reappraisal, problem-solving, and seeking social support, thereby reducing the likelihood of depression [33]. In contrast, individuals with poor emotion regulation skills are more likely to rely on avoidance, suppression, rumination, or impulsive discharge, which increases vulnerability to depressive symptoms [34]. Excessive screen time may further weaken opportunities for real-world interpersonal interaction and active emotional processing, leading students to rely more heavily on immediate and fragmented digital stimulation to regulate negative affect [35]. According to the Process Model of Emotion Regulation (PMER), individuals modulate emotions through strategies such as cognitive reappraisal and expressive suppression, which exert distinct effects on mental health [36]. On the one hand, prolonged screen use may reduce real interpersonal interaction and opportunities for authentic emotional expression, limiting practice in adaptive emotion regulation [37]. On the other hand, the immediacy, fragmentation, and emotionally salient content of online environments may foster reliance on short-term external stimulation rather than stable, internally driven regulation strategies [38]. Accordingly, emotion regulation is expected to mediate the relationship between screen time and depression. Hypothesis 3 therefore proposes that emotion regulation mediates the association between screen time and depression.

rom a mechanistic perspective, screen time, sleep quality, and emotion regulation are unlikely to operate independently; rather, they may form a sequential causal pathway. Specifically, excessive screen exposure may first impair sleep quality, leading to delayed sleep onset, insufficient sleep, and reduced daytime functioning. Subsequently, poor sleep quality may weaken cognitive control, emotional recovery, and stress regulation capacities, thereby reducing emotion regulation ability. When individuals are unable to effectively identify and regulate negative emotions, such emotions may accumulate over time and ultimately manifest as elevated depressive symptoms. In particular, excessive screen use—especially before bedtime—may delay sleep through blue light exposure, information overload, social comparison, and immersive entertainment, thereby reducing sleep quality. Impaired sleep further weakens attentional control, cognitive reappraisal, and emotion regulation capacity, making it more difficult for individuals to manage stress and negative affect, ultimately increasing depressive symptoms [39]. Sleep is a critical psychological resource that supports emotional and cognitive recovery, whereas sleep deprivation undermines these functions and leads to emotion regulation failure [40].Reduced sleep quality significantly impairs emotion regulation capacity and increases negative emotional reactivity [41], Accordingly, screen time may first affect sleep quality, then weaken emotion regulation ability, and ultimately increase depression, forming a sequential pathway. Problematic screen use is thus associated not only with sleep disturbances but also with poorer mental health outcomes [42]. Therefore, Hypothesis 4 proposes a serial mediation effect of sleep quality and emotion regulation in the relationship between screen time and depression, such that screen time increases depression by first reducing sleep quality and subsequently weakening emotion regulation capacity.

A substantial body of empirical research has reported a significant positive association between screen time and depressive symptoms. However, interpreting this association as a simple linear and causal relationship is methodologically limited and inconsistent with current scientific consensus. Recent systematic reviews, large-scale cohort studies, and meta-analyses indicate that the relationship between screen time and mental health is highly complex, nonlinear, and heterogeneous, with generally small effect sizes that are substantially moderated and mediated by multiple contextual and individual factors [43–45], Most existing studies remain at the level of correlational description, and systematic evidence on multistage, cascading mechanisms remains scarce [22, 39]. Sleep quality has been widely identified as a key psychophysiological pathway linking screen exposure and mental health outcomes. Increased screen time not only reduces sleep duration but also disrupts circadian rhythms via blue light exposure and increases cognitive and emotional arousal, thereby reducing sleep efficiency and impairing emotion regulation, ultimately elevating risks of anxiety, depression, and behavioral problems [42, 46, 47]. Nevertheless, most prior studies have examined only single-step mediation effects of sleep quality, without further exploring whether impaired sleep subsequently weakens emotion regulation capacity and thereby amplifies depression risk in a cascading manner [48]. Emotion regulation deficits have been consistently identified as robust predictors of depressive symptoms. Evidence indicates that such deficits independently predict the severity and progression of depression beyond the effects of life stressors and social support [49–51]. However, empirical studies directly examining emotion regulation as a mediator between screen time and depression remain limited, particularly in Chinese college student populations [52].

Grounded in MDT as the primary theoretical framework and complemented by SQ-ER and PMER, this study proposes a serial mediation model of “screen time → sleep quality → emotion regulation → depression” to systematically examine the association between overall screen exposure and depressive symptoms among Chinese college students and its underlying hierarchical mechanisms. Based on the literature review, the following hypotheses are proposed:

H1: Screen time positively predicts depression among college students.

H2: Sleep quality mediates the relationship between screen time and depression.

H3: Emotion regulation mediates the relationship between screen time and depression.

H4: Sleep quality and emotion regulation jointly exert a serial mediation effect between screen time and depression, such that screen time increases depression via reduced sleep quality and weakened emotion regulation.

This study provides a stratified and mechanism-informed theoretical basis for developing targeted interventions in digital health education, sleep hygiene promotion, and emotion regulation training in university settings. **Figure 1** illustrates the proposed hypothesized model.

**Figure 1.**
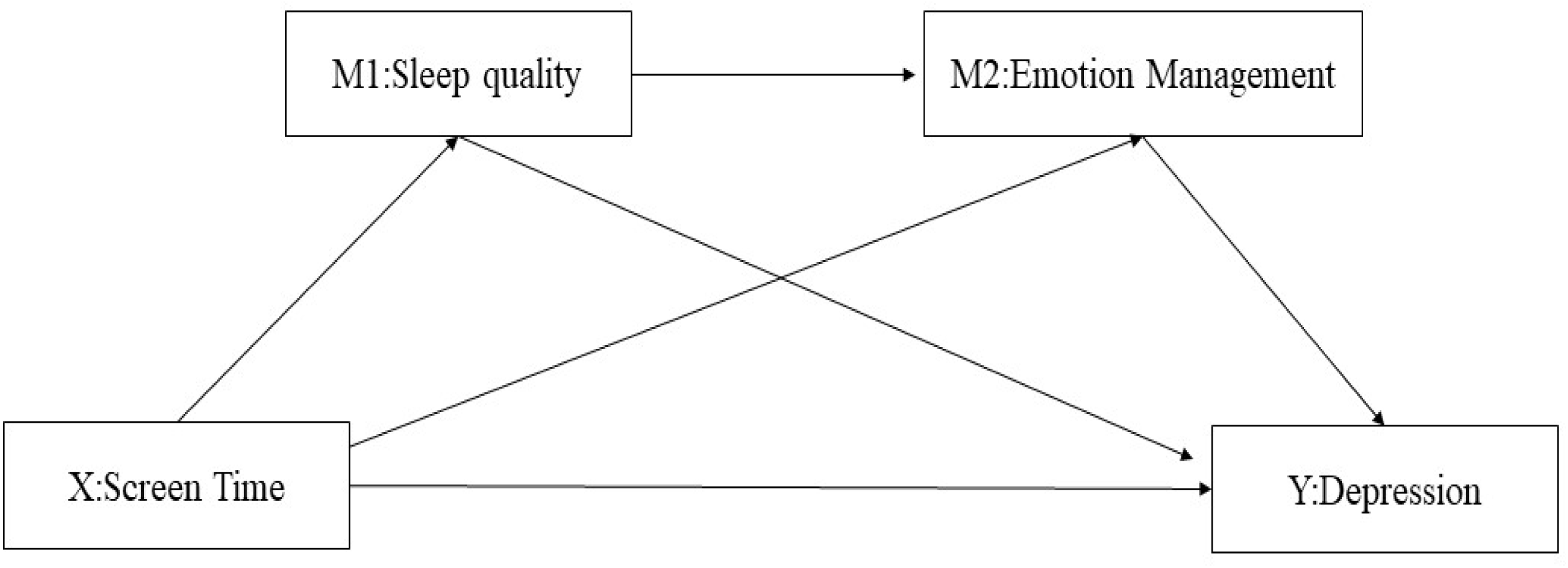
The Hypothetical Model

## 1 Participants and Methods

### 1.1 Study Population and Sampling

The data used in this study were derived from the 2024 China Physical Activity and Health Longitudinal Survey of College Students (CPAHLS-CS). The CPAHLS-CS aims to establish a nationally representative, high-resolution, individual-level database of behavioral and health-related indicators among Chinese college students, with a particular focus on physical activity and physical and mental health. The survey was designed to advance empirical research on the relationship between physical activity and health among college students in China and to facilitate interdisciplinary investigations in this field [53–63]. A stratified, cluster-based, multistage sampling strategy was adopted. Sampling procedures considered institutional type, geographical distribution, and regional socioeconomic development. Questionnaire data were collected at the level of administrative classes. The data retrieval date is November 16, 2024. The sampling frame for general higher education institutions was based on the Ministry of Education’s National List of General Higher Education Institutions, updated as of 20 June 2024.

Data quality control was conducted according to the following exclusion criteria: (1) questionnaires from institutions whose full names could not be identified were removed; (2) questionnaires containing identical responses across at least 21 consecutive items were excluded; and (3) given that the average completion time was 6 min 12 s, questionnaires with completion times falling within the lowest 0.5% or highest 0.5% of the distribution were excluded [64]].After data screening, 10,999 valid questionnaires were retained for analysis.

Detailed demographic and sample characteristics are presented in **Table 1**.

**Table 1.** Sample Distribution Table.

|  | Type | n | Percentage % |
| --- | --- | --- | --- |
| **Gender** | Male student | 4161 | 37.8 |
|  | Female student | 6838 | 62.2 |
| **Grade** | Freshman | 6855 | 66.4 |
|  | Sophomore | 3653 | 26.2 |
|  | Junior | 664 | 6.0 |
|  | Senior | 156 | 1.4 |
| **Total** |  | 10999 | 100.0 |

### 1.2 Measuring Tools

(1) Sociological Population Information

Gender and academic year were included as covariates in the present study, as both variables are closely associated with the physical and mental health of college students. Previous research has shown that, with respect to gender, female college students may be more vulnerable than their male counterparts to depressive and anxiety symptoms, which may in turn contribute to poorer mental health outcomes [65]. In terms of academic year, lower-year students, particularly those who have recently entered university, may experience rapid psychological adjustment and greater emotional fluctuations, thereby increasing their susceptibility to mental health difficulties [66]. By controlling for gender and academic year, the present study was able to more accurately examine the association between screen time and depression among college students.

(2) Screen Time

Screen time was assessed by asking participants to report their average daily duration of screen use across all electronic devices, including smartphones, computers, tablets, and televisions. A single-item question was used: “On average, how many hours per day do you spend using screens?” Based on distributional characteristics reported in previous literature and clinically relevant thresholds [67], daily screen time was categorized into three ordered groups: (1) ≤3 h/day; (2) >3 to ≤8 h/day; and (3) >8 h/day. These categories were coded as 1, 2, and 3, respectively, representing low, moderate, and high levels of screen exposure.

(3) Depression level

Depressive symptoms were assessed using the short form of the Center for Epidemiologic Studies Depression Scale–Revised (CESD-R) [68]. The original scale was developed by Richard Radloff in 1977 and was subsequently revised into a shortened version. The scale comprises two dimensions, self-acceptance and self-evaluation, with a total of nine items. Each item is rated on a 4-point scale ranging from 0 (“not at all”) to 3 (“almost every day”), with higher scores indicating more severe depressive symptoms. Based on the 80th and 95th percentiles of score distributions in Chinese age-specific populations, depressive risk was classified as follows: scores of 0–9 indicated no risk of depression, scores of 10–16 indicated mild risk of depression, and scores of 17–27 indicated severe risk of depression. The scale has demonstrated good internal consistency, with a Cronbach’s α coefficient of 0.85. In terms of validity, it showed a correlation coefficient of 0.76 with the Beck Depression Inventory (BDI), indicating satisfactory reliability and validity [68]. Therefore, the scale provides an appropriate and effective measure for assessing depressive symptom levels in the present study.

(4) Sleep quality

Sleep quality was assessed using the Pittsburgh Sleep Quality Index (PSQI), developed by Buysse and colleagues in 1989 [69]. The PSQI is a self-report instrument, with lower scores indicating better sleep quality. The scale demonstrated excellent test–retest reliability, with a reliability coefficient of 0.994. Confirmatory factor analysis further supported its construct validity, with satisfactory model fit indices, including CFI = 0.980 (>0.90), GFI = 0.970 (>0.80), and RMSEA = 0.009 (<0.08). These findings indicate that the PSQI has good reliability and validity and is an appropriate instrument for assessing sleep quality in the present study [69].

(5) Emotional management

Emotion regulation ability was assessed using the Emotional Intelligence Scale (EIS), developed by Schutte and colleagues based on the theoretical framework of Mayer and Salovey [70].The Chinese version of the scale was translated and revised by Caikang Wang at South China Normal University. The scale consists of 33 items rated on a 5-point Likert scale, ranging from 1 (“strongly disagree”) to 5 (“strongly agree”). Items 5, 28, and 33 are reverse-scored. Higher total scores indicate higher levels of emotional intelligence and stronger emotion regulation ability. The scale has demonstrated good validity, with an overall validity coefficient of 0.83, indicating that it can effectively and accurately assess emotion regulation ability among college students.

### 1.3 Statistical Methods

Data processing and statistical analyses were conducted primarily using SPSS 26.0 and Microsoft Excel. The analytical procedure comprised several key steps. First, data collected through the Wenjuanxing platform were initially screened and processed in Excel, including the rechecking or removal of incomplete and abnormal responses. Second, Harman’s single-factor test was performed to assess potential common method bias. The results showed that the largest unrotated factor accounted for 33.577% of the total variance, which was below the commonly used threshold of 40% [71], indicating that common method bias was not a serious concern in this study. Third, descriptive and inferential analyses were conducted on the student data. Chi-square tests were used to examine differences in depression levels across gender and academic year groups. Effect size was evaluated using η², with values of 0.01, 0.06, and 0.14 representing small, medium, and large effects, respectively [72]. Fourth, Pearson correlation analyses were performed to examine the associations among screen time, sleep quality, emotion regulation, and depression. Finally, regression analyses were conducted to test the mediation effects. The PROCESS macro for SPSS was used for further mediation analysis, with Model 6 selected to examine the serial mediation model. The confidence interval was set at 95%, and 5,000 bootstrap samples were used to estimate the indirect effects.

## 2 Result

### 2.1 Descriptive Result Analysis

As shown in **Table 2**, the mean screen time score among college students was 2.07 ± 0.53. Female students reported slightly higher screen time than male students (2.09 vs. 2.05), and the difference was statistically significant (F = 11.30, p < 0.001, η² = 0.001). Significant differences were also observed across academic years (F = 26.962, p < 0.001, η² = 0.007), with third-and fourth-year students scoring higher (2.22 and 2.21, respectively) than first-and second-year students (2.05 and 2.10, respectively). The effect of academic year was larger than that of gender. The mean depression score was 15.79 ± 5.04. Male students reported higher depression scores than female students (16.08 vs. 15.62), and this gender difference was statistically significant (F = 21.80, p < 0.001, η² = 0.002). Depression scores also differed significantly by academic year (F = 45.55, p < 0.001, η² = 0.012), with the highest score observed among fourth-year students (16.91) and the lowest among first-year students (15.40). The effect of academic year was greater than that of gender. The mean sleep quality score was 2.06 ± 0.80. Male students had higher sleep quality scores than female students (2.11 vs. 2.03), indicating poorer sleep quality among males, and the gender difference was statistically significant (F = 25.99, p < 0.001, η² = 0.002). Significant differences were also found across academic years (F = 34.48, p < 0.001, η² = 0.009), with fourth-year students showing the highest scores (2.33) and first-year students the lowest scores (2.01). The effect of academic year was again larger than that of gender. The mean emotion regulation score was 29.62 ± 4.40. No significant gender difference was found (F = 0.56, p = 0.454, η² < 0.001). Although emotion regulation differed significantly across academic years (F = 3.85, p = 0.009, η² = 0.001), the effect size was extremely small, and mean scores were broadly comparable across academic years, ranging from 29.10 to 30.04.

**Table 2.**
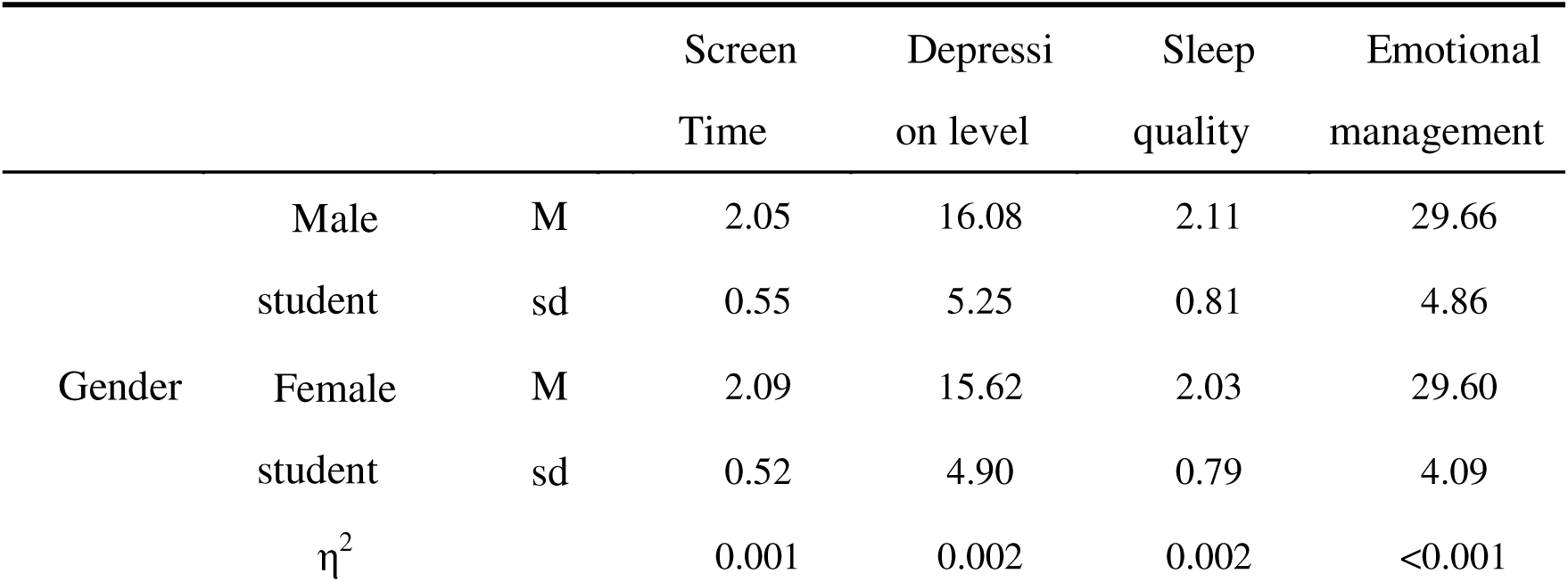
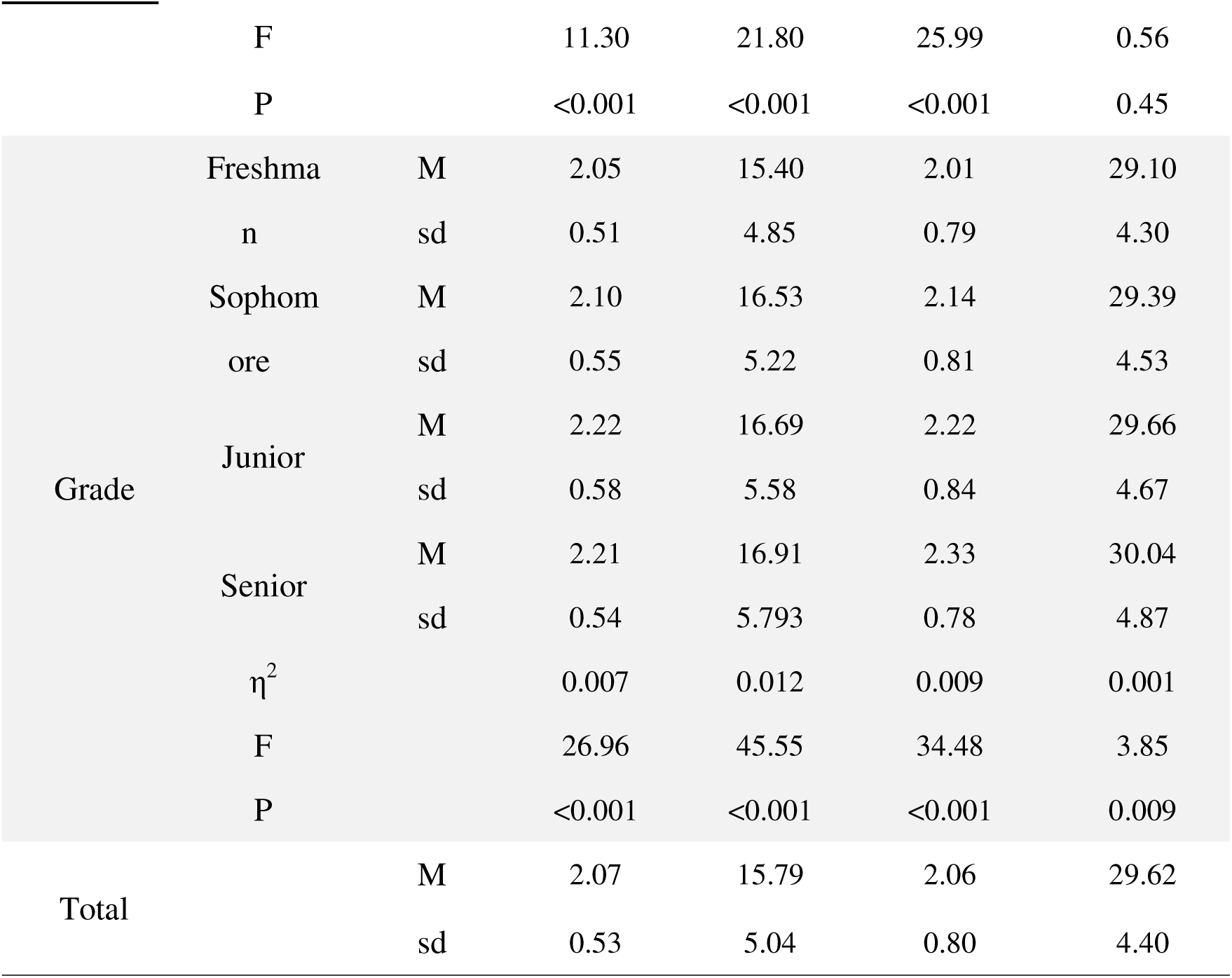
Descriptive Statistics of the Variables.

### 2.2 Correlation Analysis

As shown in **Figure 2**, screen time was positively correlated with depressive symptom severity (r = 0.16) and sleep-quality scores (r = 0.15), but negatively correlated with emotion regulation (r = −0.13). Depressive symptom severity was positively associated with sleep-quality scores (r = 0.31) and negatively associated with emotion regulation (r = −0.42). In addition, sleep-quality scores were negatively correlated with emotion regulation (r = −0.21).

**Figure 2.**
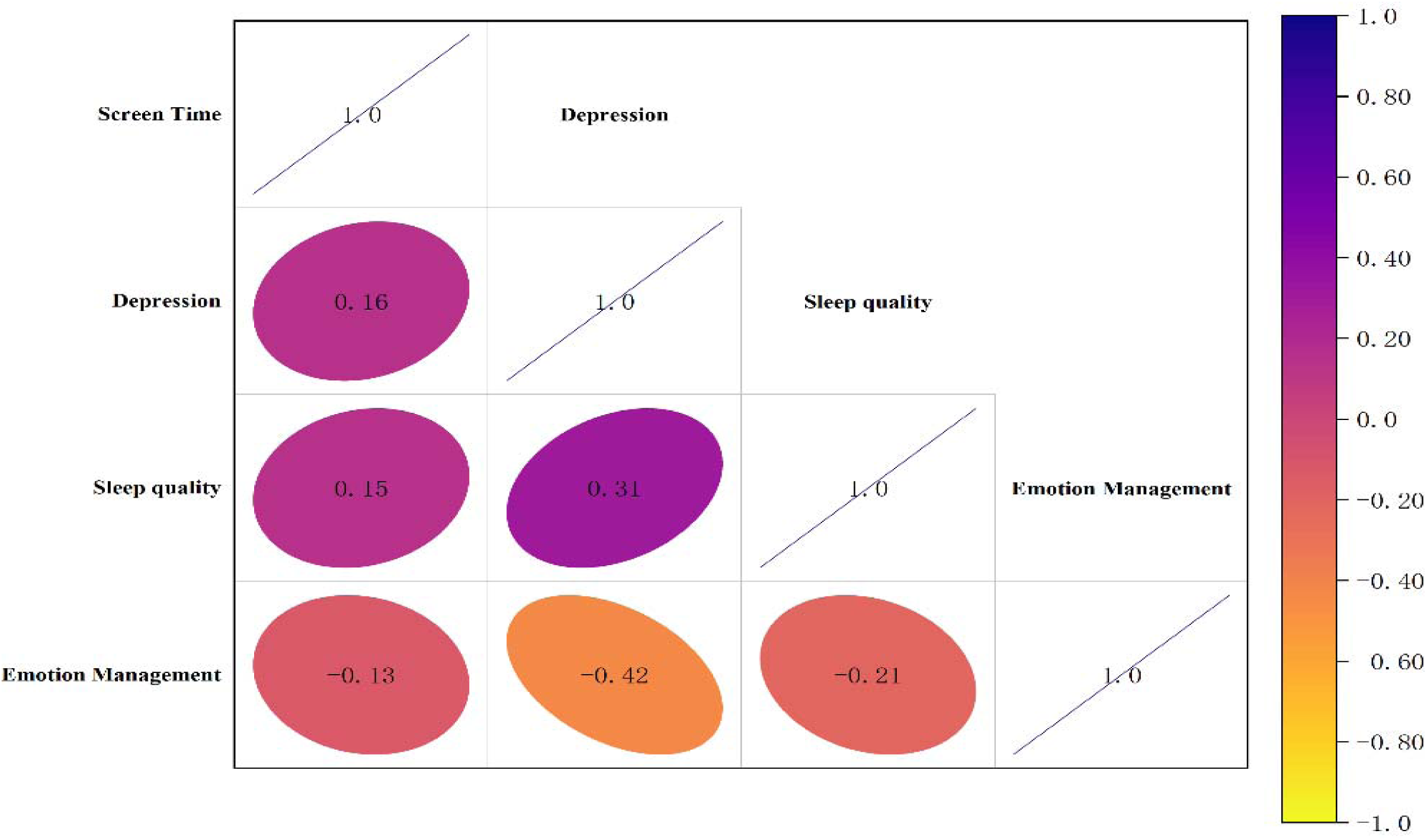
Correlation Analysis Heatmap

### 2.3 Regression Analysis

Before conducting the formal model tests, scatterplots were used to visually inspect the linear relationships among the study variables. As shown in **Figure 3**, the regression scatterplots of the core variables depict the simple linear relationship between screen time and depressive symptom severity among college students, together with the regression trends for the predictor variables in the model. The scatter distributions and fitted lines provide intuitive evidence regarding the direction and magnitude of these linear associations, which is consistent with the regression model results and supports the subsequent mediation analysis.

**Figure 3.**
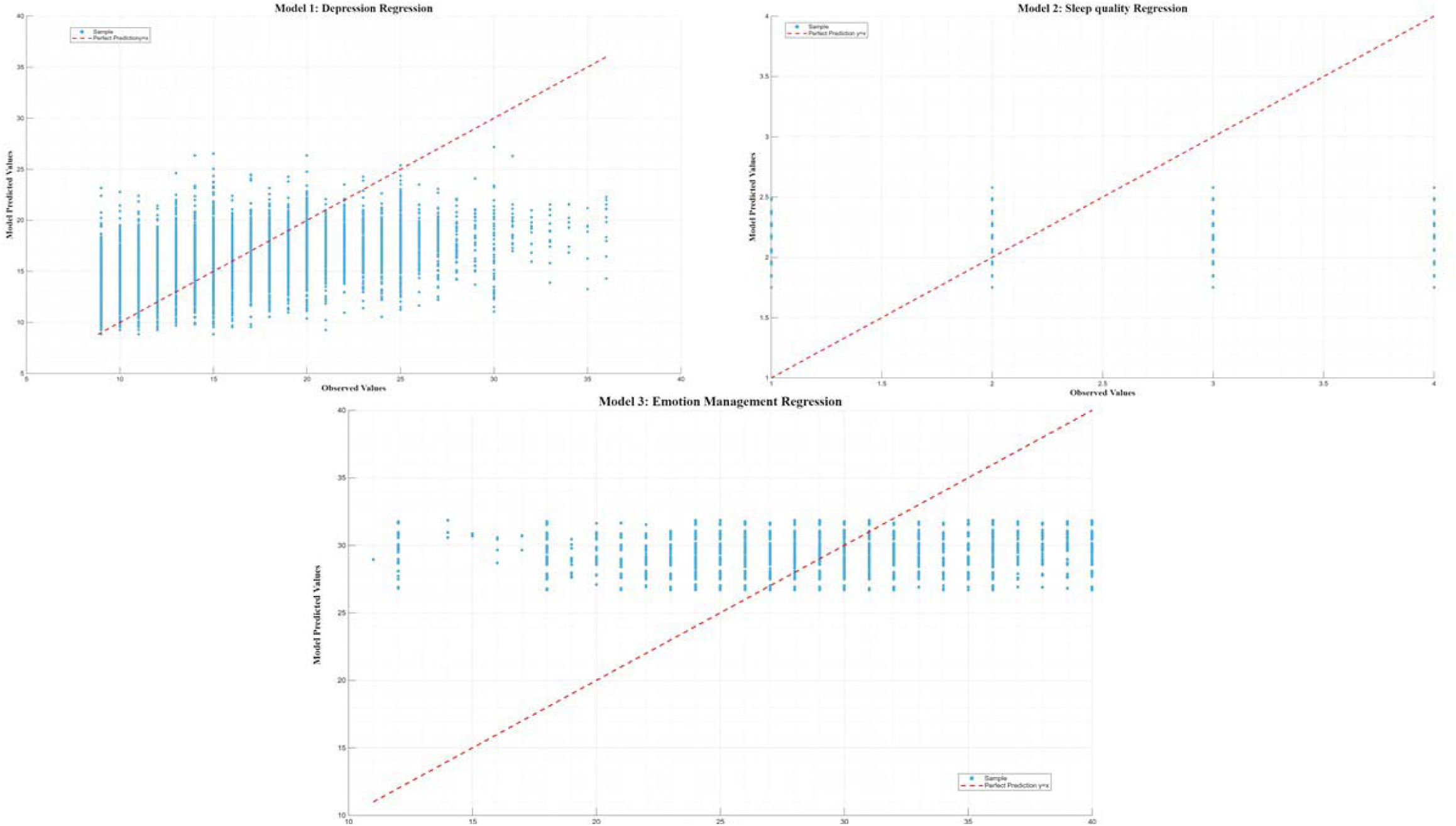
Regression Scatter Plots

Multiple regression analyses were conducted using SPSS 26.0 to examine the independent predictive effects of screen time on depressive symptom severity, sleep quality, and emotion regulation, while controlling for sex and grade level. Where appropriate, sleep quality and emotion regulation were further included as covariates to account for their potential confounding effects. As shown in **Table 3**, after adjustment for relevant covariates, screen time significantly and positively predicted depressive symptom severity (β = 0.712, P < 0.001) and sleep quality scores (β = 0.217, P < 0.001), while significantly and negatively predicting emotion regulation (β = −0.085, P < 0.001). Sleep quality scores (β = 1.318, P < 0.001) and emotion regulation (β = −0.424, P < 0.001) also emerged as independent predictors of depressive symptom severity. In addition, sex and grade level showed significant predictive effects across all regression models (P < 0.01).

**Table 3.**
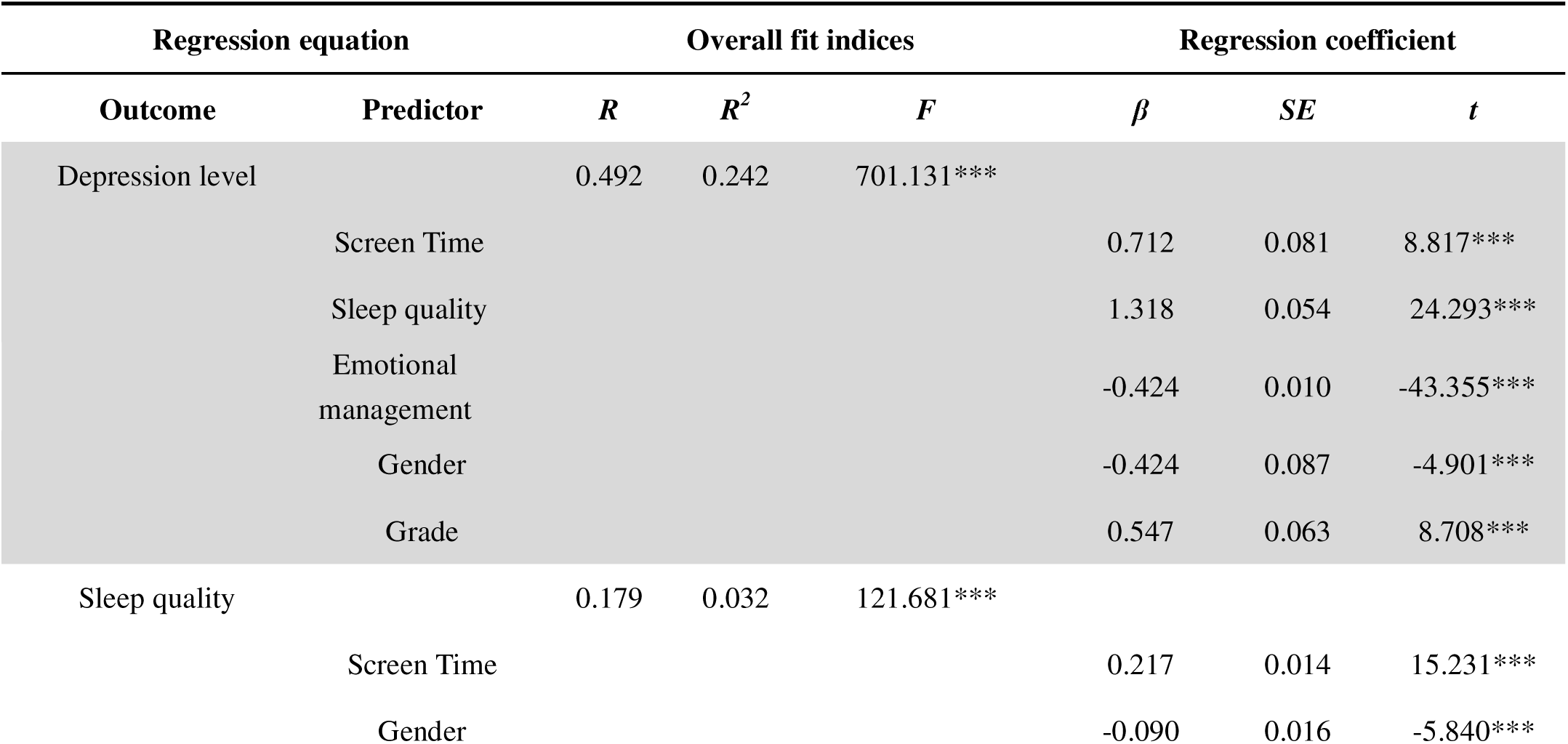
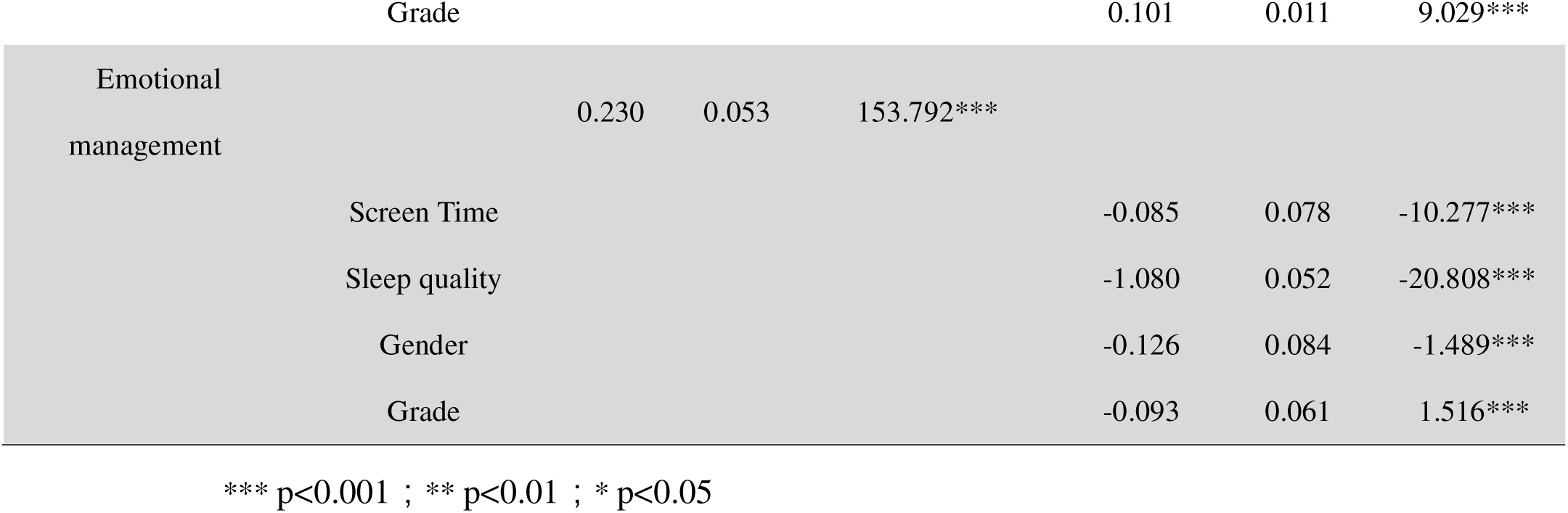
Summary of Regression Analysis with Confounding Factors Excluded.

### 2.4 Mediation Effect Test

As presented in **Table 4**, screen time exerted a significant total effect on depressive symptom severity (effect = 1.439, 95% CI [1.263, 1.614]). The direct effect remained significant (effect = 0.712, 95% CI [0.554, 0.870]), representing 49.48% of the total effect, while the overall indirect effect was also significant (effect = 0.727, 95% CI [0.633, 0.819]), representing 50.52% of the total effect. The 95% confidence intervals for all three mediating pathways excluded zero, indicating significant mediation effects. Specifically, sleep quality significantly mediated the association between screen time and depressive symptom severity (effect = 0.286, 95% CI [0.239, 0.332]; proportion mediated = 19.87%), as did emotion regulation (effect = 0.341, 95% CI [0.269, 0.416]; proportion mediated = 23.70%). In addition, the sequential indirect pathway from screen time to depressive symptom severity through sleep quality and emotion regulation was significant (effect = 0.100, 95% CI [0.082, 0.117]; proportion mediated = 6.95%).

**Table 4.**
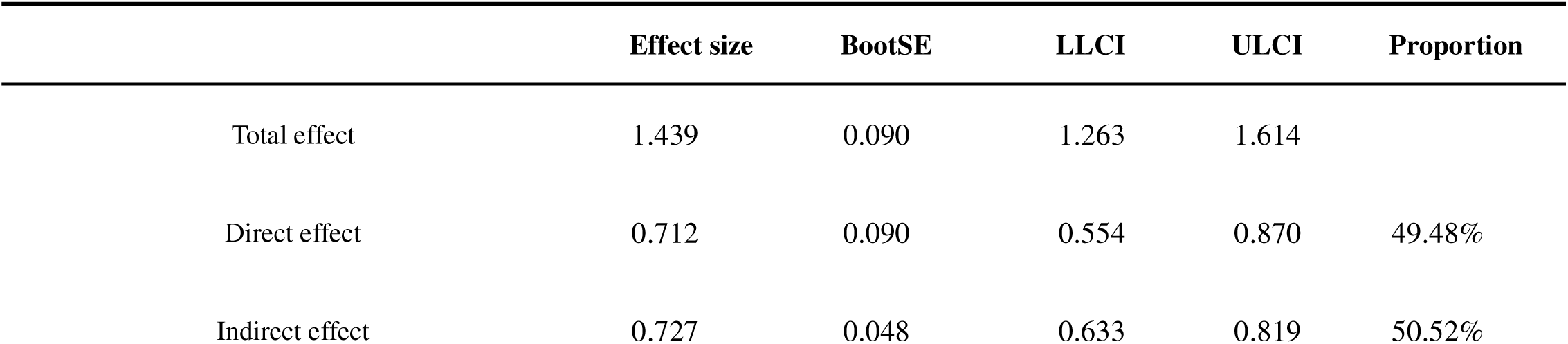
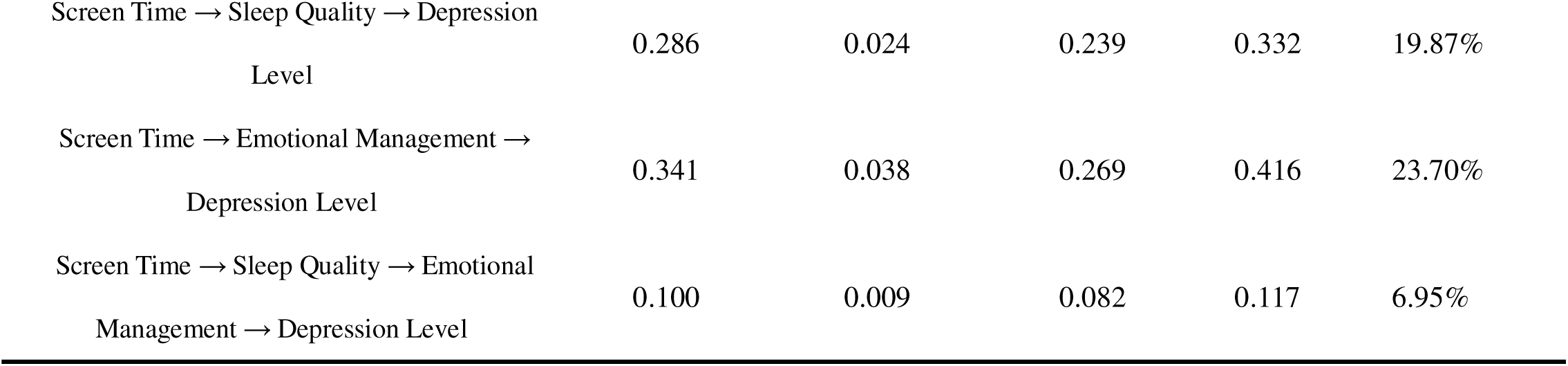
Summary of Mediation Effect Analysis.

|  | Effect size | BootSE | LLCI | ULCI | Proportion |
| --- | --- | --- | --- | --- | --- |
| Total effect | 1.439 | 0.090 | 1.263 | 1.614 |  |
| Direct effect | 0.712 | 0.090 | 0.554 | 0.870 | 49.48% |
| Indirect effect | 0.727 | 0.048 | 0.633 | 0.819 | 50.52% |
| Screen Time → Sleep Quality → Depression<br>Level | 0.286 | 0.024 | 0.239 | 0.332 | 19.87% |
| Screen Time → Emotional Management →<br>Depression Level | 0.341 | 0.038 | 0.269 | 0.416 | 23.70% |
| Screen Time → Sleep Quality → Emotional<br>Management → Depression Level | 0.100 | 0.009 | 0.082 | 0.117 | 6.95% |

Higher sleep quality scores indicate poorer sleep quality, higher emotion regulation scores indicate stronger emotion regulation capacity, and higher depression scores indicate more severe depressive symptoms. The path coefficient from screen time to depressive symptoms was 0.712 (P < 0.001), indicating that screen time had a significant positive effect on the severity of depressive symptoms. The path coefficient from screen time to emotion regulation was −0.085 (P < 0.001), suggesting a significant negative effect of screen time on emotion regulation. The direct path coefficient from screen time to sleep quality was 0.217 (P < 0.001), indicating that greater screen time was significantly associated with poorer sleep quality. The path coefficient from sleep quality to depressive symptoms was 1.318 (P < 0.001), suggesting that poorer sleep quality significantly contributed to higher levels of depressive symptoms. The path coefficient from sleep quality to emotion regulation was −0.108 (P < 0.001), indicating that poorer sleep quality exerted a significant negative effect on emotion regulation. In addition, the path coefficient from emotion regulation to depressive symptoms was −0.424 (P < 0.001), indicating that stronger emotion regulation was significantly associated with lower levels of depressive symptoms. Taken together, these findings suggest that longer screen time may impair sleep quality and weaken emotion regulation, thereby contributing to elevated depressive symptoms. These pathways collectively constitute the mediation model linking screen time to depressive symptoms, as illustrated in **Figure 4**.

**Figure 4.**
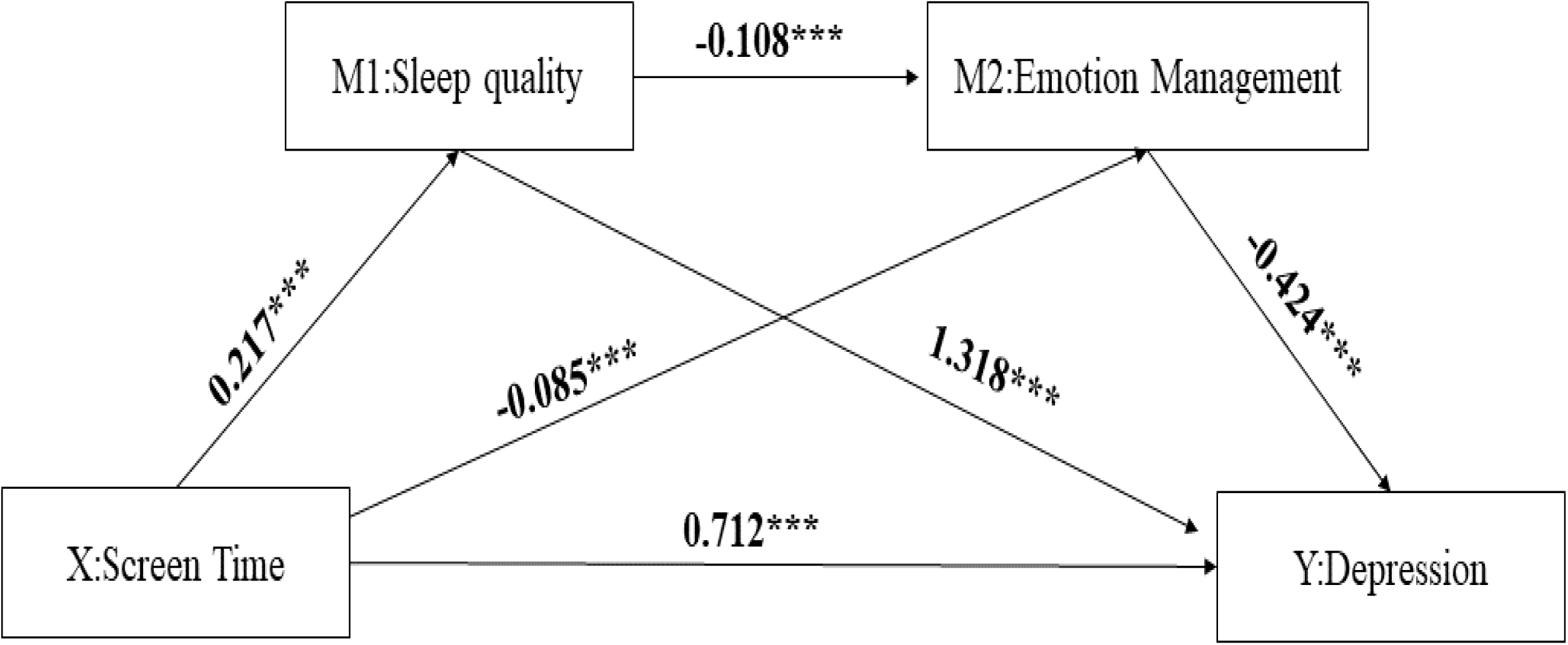
Mediating Effect Diagram

**Figure 5** presents the visualization of the standardized path coefficients among screen time, sleep quality, emotion regulation, and depression. As shown in the figure, screen time significantly and positively predicted poorer sleep quality and depression, while significantly and negatively predicting emotion regulation. Poorer sleep quality significantly and positively predicted depression and significantly and negatively predicted emotion regulation. In addition, emotion regulation exerted a significant negative predictive effect on depression. All path coefficients reached statistical significance, and the visualization results were consistent with the findings obtained from the model regression analyses.

**Figure 5.**
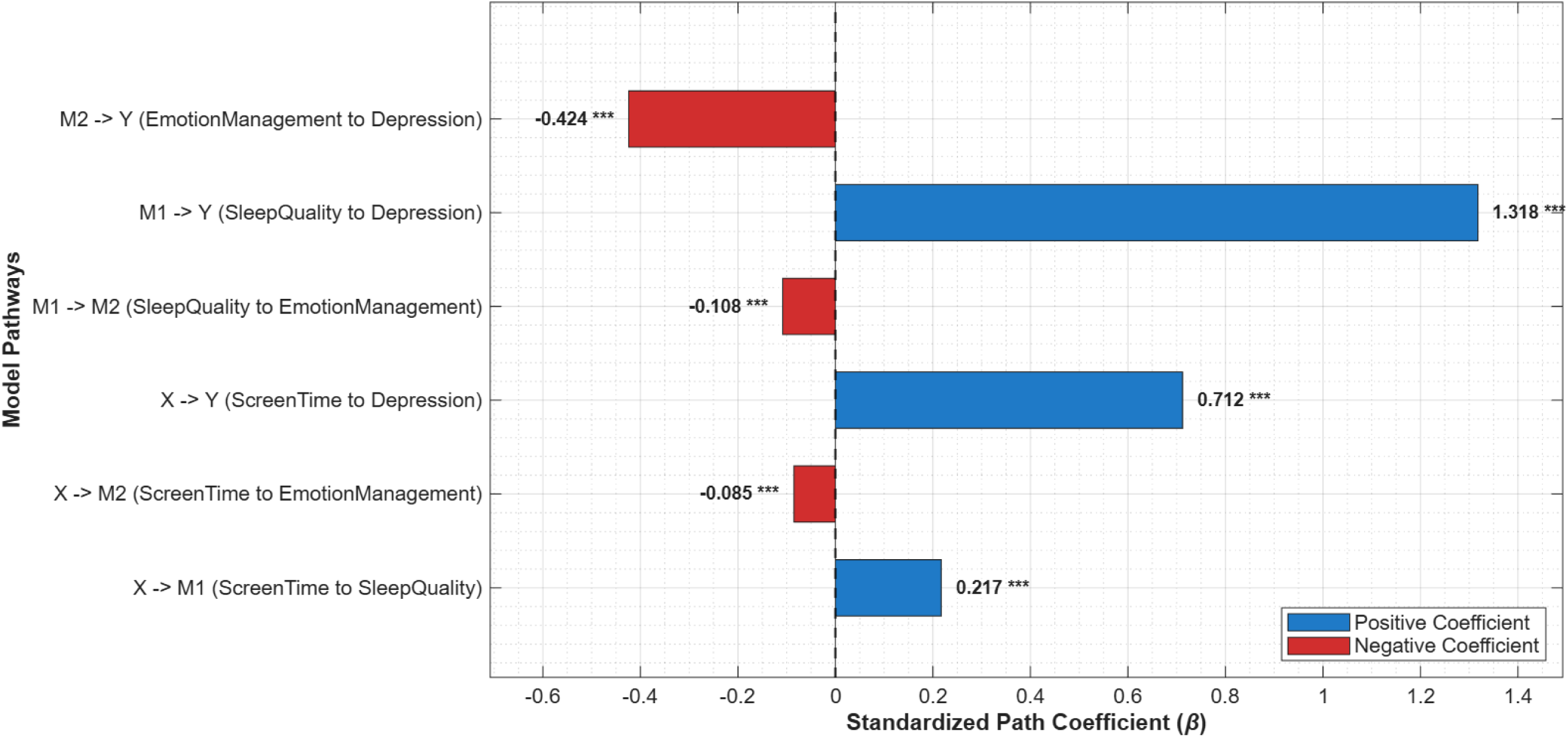
Visualization of Regression Coefficients for Mediating and Moderating Paths

## 3 Discussion

Drawing on media displacement theory, the sleep–emotion regulation model, and emotion regulation theory, this study systematically examined the association between screen time and depression among Chinese university students, with a particular focus on the mediating roles of poorer sleep quality and emotion regulation. The findings showed that screen time was significantly and positively associated with depression, which is broadly consistent with previous evidence and further supports the view that digital media use is closely linked to mental health outcomes. Beyond demonstrating the independent mediating effects of poorer sleep quality and emotion regulation, this study further identified a sequential pathway through which screen time may be associated with depression via poorer sleep quality and impaired emotion regulation. This finding extends prior research that has primarily focused on single mediating mechanisms and provides a more integrated explanatory framework for understanding mental health problems among university students.

The results showed that the total effect of screen time on depression was 1.439, with a direct effect of 0.712 and an indirect effect of 0.727. These findings suggest that screen time is not only directly associated with depressive symptoms among university students but may also be indirectly linked to depression through poorer sleep quality and emotion regulation. Specifically, longer screen time may be associated with poorer sleep quality and reduced emotion regulation capacity, both of which may further contribute to higher levels of depression among university students. Overall, this study enriches the explanatory framework for understanding the relationship between screen time and mental health in university students and provides empirical evidence for the development of digital health education, sleep health promotion, and emotion regulation interventions in higher education settings.

### 3.1 Direct Effect of Screen Time on Depression Levels

This study first confirmed the positive predictive effect of screen time on depressive symptoms among university students (β = 0.712, P < 0.001), thereby supporting Hypothesis H1. This finding is consistent with evidence from a study of U.S. adults, which showed that individuals with longer daily screen time reported higher levels of depression than those with shorter screen time[73]. In addition, a meta-analysis comprising multiple cohort studies reported a significant statistical association between increased screen time and elevated risk of depression[45]. These findings suggest that, even after accounting for potential mediating variables, screen time itself retains substantial explanatory power in relation to depressive symptoms.

However, the direct effect of screen time has not been equally pronounced across all studies. Using large-scale data from the 2018 National Survey of Children’s Health in the United States, Leung et al. found that sleep duration did not mediate the associations between screen time and depression or anxiety among adolescents[74]. In contrast, the direct effect remained significant in the present study. This discrepancy may be partly attributable to sample characteristics. Our sample included students from the first to the fourth year of university, representing a relatively broad range of academic stages. Notably, fourth-year students reported the longest screen time and the highest levels of depressive symptoms, suggesting that grade level may have amplified the observed direct effect. Moreover, previous studies have indicated that the type of screen use is a critical moderating factor: educational screen use tends to show weaker, or even non-significant, associations with depression[75], whereas social and recreational screen use predicts poorer mental health outcomes[76]. Because the present study assessed overall screen time without distinguishing between types of use, the observed direct effect may have been driven primarily by non-educational screen activities. This also suggests that the direct effect estimated in this study should be interpreted as a relatively coarse-grained estimate. Future research that differentiates screen use by content or purpose may reveal substantial heterogeneity in the strength of the direct effect across different forms of screen engagement.

In summary, this study confirmed a direct positive association between screen time and depressive symptoms. Nevertheless, the magnitude of this association may vary depending on measurement approaches, the academic-year composition of the sample, and the specific content or purpose of screen use. The presence of a significant direct effect provides an important foundation for further examination of the mediating mechanisms linking screen time to depressive symptoms.

### 3.2 The Mediating Effect of Sleep Quality

The present findings revealed that sleep quality significantly mediated the association between screen time and depressive symptoms, thereby supporting Hypothesis H2. Correlational analyses further showed that screen time was positively associated with sleep quality scores and that sleep quality scores were positively associated with depressive symptom levels. Given that higher scores on the sleep quality measure used in this study indicate poorer sleep quality, these results suggest that university students who spend more time on screens may experience poorer sleep, which in turn is associated with more severe depressive symptoms. This finding is consistent with previous evidence indicating that prolonged screen exposure, particularly the use of electronic devices before bedtime, may disrupt sleep rhythms and impair sleep restoration by delaying sleep onset and increasing cognitive arousal[77].

This result is also consistent with the core proposition of MDT. Among university students, screen use includes not only academic activities but also non-academic activities such as social networking, entertainment, and information browsing[78]. As screen time increases, it may displace time that would otherwise be allocated to rest, physical activity, and face-to-face social interaction[79]. This displacement may be especially detrimental when electronic devices are used before bedtime, as such use may delay sleep onset, heighten psychological arousal, and weaken the restorative function of sleep[80]. Because sleep is a fundamental basis for maintaining cognitive functioning[81] and emotional stability[82], poor sleep quality may reduce individuals’ capacity to recover from daily stressors, thereby increasing the risk of depressive symptoms[30].

From the perspective of the SQ-ER framework, sleep quality is not merely a physiological recovery process, but also a critical condition for the effective operation of emotion regulation systems[83]. University students are situated at a developmental stage characterized by multiple concurrent demands, including academic pressure, interpersonal adjustment, and preparation for future employment. Prolonged screen exposure combined with declining sleep quality may therefore undermine emotional stability, intensify negative cognition, and weaken psychological recovery capacity. In the present study, the significant indirect pathway from screen time to depressive symptoms through sleep quality indicates that sleep quality may constitute an important psychophysiological mechanism linking screen time to depressive symptoms. Accordingly, university-based interventions aimed at managing students’ screen time should not focus solely on reducing the duration of electronic device use. They should also incorporate sleep hygiene education, encourage students to limit prolonged screen exposure before bedtime, and promote regular sleep–wake schedules, thereby helping to reduce the risk of depressive symptoms.

### 3.3 The Mediating Effect of Emotion Management

The present study found that emotion management significantly mediated the association between screen time and depressive symptoms, thereby supporting Hypothesis H3. Specifically, screen time negatively predicted emotion management, whereas emotion management negatively predicted depressive symptom levels. These findings suggest that university students with longer screen time may exhibit weaker emotion management abilities, and that lower emotion management capacity is associated with higher levels of depressive symptoms.

This result is consistent with the PMER framework. Individuals with stronger emotion management abilities are generally better able to identify, understand, and regulate their emotional experiences[84]. When confronted with academic stress, interpersonal conflict, and uncertainty about the future, they may be more likely to adopt adaptive strategies such as cognitive reappraisal, problem solving, and seeking social support[85]. In contrast, university students with insufficient emotion management abilities may be more vulnerable to rumination, avoidance, or emotional suppression. These maladaptive responses may contribute to the persistent accumulation of negative affect, thereby increasing the risk of depressive symptoms[86].

The influence of screen time on emotion management may operate through multiple mechanisms. On the one hand, prolonged screen use may reduce opportunities for university students to engage in real-world interpersonal interaction and emotional expression, thereby weakening the process through which they learn and practice emotion regulation strategies in authentic social relationships[87]. On the other hand, social comparison, fragmented stimulation, and negative content on social media may increase individuals’ emotional burden[88]. As a result, students may become more reliant on immediate online stimulation to alleviate negative emotions, rather than developing stable and proactive emotion management strategies[89]. It should be noted, however, that the cross-sectional design of the present study precludes causal inference; therefore, the findings cannot establish that screen time necessarily leads to poorer emotion management. Nevertheless, the results suggest that emotion management may represent an important psychological mechanism underlying the association between screen time and depressive symptoms.

Accordingly, psychological health education in universities may benefit from integrating emotion management training with education on digital media use[90]. Through group counseling, emotion recognition training, stress management programs, and cognitive reappraisal training, universities can help students develop more adaptive emotion regulation strategies and reduce passive reliance on screen-based stimulation, thereby lowering the risk of depressive symptoms[91].

### 3.4 The Chain Mediating Effect of Sleep Quality and Emotional Management

The present study further revealed a significant chain-mediating effect of sleep quality and emotion management in the association between screen time and depressive symptoms, thereby supporting Hypothesis H4. Specifically, screen time was not only associated with depressive symptoms through the independent mediating roles of sleep quality and emotion management, but was also linked to depressive symptom levels through the sequential pathway of “screen time → sleep quality → emotion management → depressive symptoms.”

This finding suggests that screen time, sleep quality, and emotion management are not isolated factors, but may operate within a progressive mechanism. Longer screen time may first be associated with poorer sleep quality among university students, manifested by difficulties initiating sleep, insufficient sleep duration, or impaired daytime functioning. Deterioration in sleep quality may subsequently compromise cognitive control, attentional regulation, and emotional recovery capacity[92], hereby weakening students’ ability to manage emotions effectively[16]. When university students have difficulty identifying and regulating negative emotions, negative affect may accumulate more persistently and ultimately be reflected in higher levels of depressive symptoms.

At the theoretical level, this sequential pathway illustrates the internal linkage among MDT, the SQ-ER framework, and PMER. MDT explains how increased screen time may displace resources otherwise allocated to sleep and real-world activities[27]. The sleep–emotion regulation model suggests that poorer sleep quality may undermine emotion regulation capacity[93]. Emotion regulation theory further explains how insufficient emotion management may increase vulnerability to depressive symptoms[94]. Together, these theoretical perspectives provide a continuous explanatory framework for understanding how screen time may be associated with university students’ mental health.

Compared with previous studies that have primarily focused on single mediating variables, the present study extends this literature by demonstrating the interconnection among multiple psychological and psychophysiological mechanisms through a chain mediation model. The results indicate that screen time may first be related to poorer sleep quality, which in turn may be associated with weaker emotion management capacity, ultimately contributing to higher levels of depressive symptoms. This integrated perspective connects media displacement theory, the sleep–emotion regulation model, and emotion regulation theory, thereby offering a more comprehensive framework for explaining the relationship between screen time and mental health[95]. In parallel, sleep hygiene education may be implemented to improve students’ overall sleep quality[96]. Building on this foundation, universities should also strengthen emotion management training through mental health courses, group counseling, or mindfulness-based interventions, thereby enhancing students’ ability to identify, express, and regulate emotions[97].

In addition, the present findings suggest that university mental health service systems may consider the pattern of “high screen time–sleep problems–emotion regulation difficulties” as a sequential risk signal during screening and intervention. Compared with monitoring a single indicator, this multifactorial assessment perspective may improve the sensitivity of early warning systems for psychological problems and enhance the precision of targeted interventions.

### 3.5 Limitations

Several limitations should be acknowledged. First, the cross-sectional design of this study allows only the identification of associations among variables and statistically inferred pathways, precluding firm causal conclusions. Future studies should employ longitudinal or experimental intervention designs to further clarify the temporal ordering and causal links among screen time, sleep quality, emotion regulation, and depressive symptoms. Second, the study relied primarily on self-report questionnaires, which may be subject to recall bias and social desirability bias. Subsequent research would benefit from incorporating objective measures, such as digital records of screen use and sleep parameters obtained from wearable devices, to enhance measurement validity. Third, screen time was examined as an overall level of use, without differentiating between academic, social, and recreational screen-based activities. Given that different forms of screen use may exert distinct effects on sleep quality and mental health, future work should adopt a more refined classification of screen-use types. Fourth, although demographic variables such as sex and grade level were controlled for, other factors—including family socioeconomic status, academic stress, physical activity, social support, and personality traits—may also influence depressive symptoms among university students. Incorporating these variables in future studies may further improve the explanatory power of the proposed model.

## 4. Conclusion

This study developed and tested a serial mediation model to systematically examine the association between screen time and depressive symptoms among Chinese university students, as well as the underlying mechanisms linking these variables. The results indicated that screen time not only had a significant direct positive effect on depressive symptoms, but also exerted indirect effects through multiple mediating pathways involving sleep quality and emotion management. Specifically, longer screen time was associated with poorer sleep quality, which in turn was linked to reduced emotion management capacity; diminished emotion management capacity was subsequently associated with higher levels of depressive symptoms. These findings suggest that the adverse impact of screen time on mental health may be transmitted through a progressive resource-depletion process characterized by impaired physiological restoration followed by weakened psychological regulatory capacity.

By proposing and validating this serial mediation model, the present study extends the theoretical framework concerning the relationship between screen time and mental health from an integrative perspective. In particular, it connects media displacement theory, the sleep–emotion regulation model, and emotion regulation theory, thereby elucidating the internal logical links among multiple psychological mechanisms. These findings further suggest that mental health interventions in higher education settings should not be confined to a single dimension. Instead, comprehensive promotion strategies that integrate screen-use management, sleep hygiene education, and emotion management training may be more effective in reducing depressive symptoms and promoting the physical and psychological well-being of university students.

## Abbreviations

BDI: Beck Depression Inventory
CESD-R: Center for Epidemiologic Studies Depression Scale–Revised
CFI: Comparative Fit Index
CPAHLS-CS: China Physical Activity and Health Longitudinal Survey of College Students
EIS: Emotional Intelligence Scale
GFI: Goodness-of-Fit Index
MDT: Media Displacement Theory
PMER: Process Model of Emotion Regulation
PSQI: Pittsburgh Sleep Quality Index
RMSEA: Root Mean Square Error of Approximation
SQ-ER: Sleep–Emotion Regulation model

## Acknowledgments

We sincerely thank all the staff and students from the participating schools and our co-operators for their assistance in data collection.

## Funding

This study was supported by the 1) Postgraduate Research & Practice Innovation Program of Jiangsu Province

## Data availability

The data that supports the findings of this study are available on request from the corresponding author, upon reasonable request.

## Declarations

### Ethics approval and consent to participate

This study was conducted in accordance with the ethical principles stated in the Declaration of Helsinki. The study protocol for this study received approval from the ethics committee at Nantong Municipal Health Commission and was documented under approval number 2024(1). Prior to commencing the formal investigations and testing, the researchers obtained informed consent from all the participants involved in the study.

### Consent for publication

Applicable.

### Competing interests

The authors declare that they have no competing interests.

## Author summary

### Authors contributions

Liang Chen completed the literature review and wrote the main manuscript text. Kai-xin Li prepared all the tables and figures. Ding-you Zhang and Bo Li completed the data analysis. Shi-hai Yu and Shan-shan Han reviewed the manuscript. Liang Chen, Sai Zhu, and Hu Lou participated in and completed tasks, including onsite investigations, data organization, and data management, in the survey.

## Notes

### Competing Interest Statement

The authors have declared no competing interest.

### Author Declarations

The study protocol for this study received approval from the ethics committee at Nantong Municipal Health Commission and was documented under approval number 2024(1).

